# Exploring the health impacts and inequalities of the new way of working: findings from a cross-sectional study

**DOI:** 10.1101/2022.01.07.22268797

**Authors:** Melda Lois Griffiths, Benjamin J Gray, Richard G Kyle, Alisha R Davies

## Abstract

**Aim:** To explore the working Welsh adult population’s ability to work from home, their preferences for the future, and the self-reported health impacts of home-working.

**Subject and Methods:** A nationally-representative household survey was undertaken across Wales (Public Health Wales’ *COVID-19, Employment and Health in Wales* study), with cross-sectional data on home-working being collected between November 2020 and January 2021 from 615 employed working-aged adults in Wales (63.7% female, 32.7% aged 50-59). Respondents were asked about their ability to work from home, their perceptions of its impact on their health and their preferences for time spent home-working in future.

**Results:** Over 50% were able to work from home, and showed a preference towards home-working to some capacity, with over a third wishing to work from home at least half the time. However, those living in the most deprived areas, in atypical employment, with high wage precarity or with limiting pre-existing conditions were less likely to report being able to work from home. Of those that could work from home, over 40% reported that it worsened their mental well-being and loneliness, and for people in poorer health, home-working negatively impacted their diet, physical activity, smoking and alcohol use. People aged 30 to 39 and those who lived alone were more likely to report wanting to spend some time working in an office/base instead of at home.

**Conclusion:** The inequity in the ability to work from home reflects underlying inequalities in Wales, with those facing the greatest insecurity (e.g. those living in most deprived areas, those with more precarious work or financial circumstances) being less able to participate in home-working. Working from home offers greater flexibility, reduces the financial and time costs associated with commuting, and protects individuals from exposure to communicable diseases. However, working from home presents an enormous challenge to preserving the mental-wellbeing of the workforce, particularly for younger individuals and those with low mental well-being. Younger respondents and those in poorer health who could work from home were also more likely to engage in health-harming behaviours, and reduce their engagement in health-protective behaviours such as eating well and moving more. Reflecting on the future, providing pathways for accessing work from home arrangements, integrating hybrid models and preparing targeted health support for at risk groups may be best suited to the working population’s preferences and needs.

## Introduction

Strict home-working requirements have been implemented by numerous administrations globally as a key strategy to slow the spread of Coronavirus (SARS-CoV-2) (Rubin et al. 2020; Arshed et al. 2020; Miles et al. 2021). This has led to one of the most striking societal transformations of the COVID-19 pandemic. Home-working, historically the privilege of a small minority of workers (Felstead et al. 2002), is now the new norm for many (Felstead and Reuschke 2020). Estimations from early on in the pandemic suggested that around 50% of Europeans worked from home to some extent as a result of the pandemic, compared to only 12% before its onset (Eurofound 2020). The COVID-19 pandemic serves as an unplanned experiment at mass scale (Kramer and Kramer 2020) – studying this transition into home-working and people’s preferences for the future is vital.

Home working can offer benefits for individuals, employers and society. For example, working from home offers greater flexibility. Having the option to work from home could make work and its associated health benefits more accessible for sub-groups that need greater flexibility e.g. those with care responsibilities or those dealing with health conditions which may make accessing work more challenging (Beatty and Joffe 2006; Waddell and Burton 2006; Holland and Collins 2018; UK Government 2021). In turn, improving the accessibility of work protects against unemployment and its negative health impacts (van Aerden et al. 2017). Home-working may also improve work-life balance and the productivity of work, while also offering protection from exposure to communicable diseases (Felstead and Reuschke 2020; Dyakova et al. 2021). On a societal level, home-working may provide an attractive opportunity to contribute to protecting the climate through reducing carbon-emissions generated by the commute (Hook et al. 2020; Bachelet et al. 2021; Beno 2021). However, the pandemic has also shed light on some of the potential negative outcomes of a more permanent adoption of work from home policies. The burden on mental health has been well-documented throughout the COVID-19 pandemic. Working from home can be isolating, negatively impacting mental well-being and levels of physical activity, and holding the potential to detrimentally impact physical health for those with inadequate resources to ensure a safe working environment at home e.g. ergonomic equipment (Dyakova et al. 2021).

Despite these potential disadvantages, there is both population and policy level evidence to suggest that the transition to home-working has been welcomed. Of those within the UK population who have worked from home during the pandemic, 88.2% wish to continue to some degree, with 47.3% wishing to work from home either often or all the time (Felstead and Reuschke 2020). Several governments have announced their support for adopting work from home policies more permanently. For example, in 2020 the German labour minister stated his intention to publish a draft law establishing the legal right to work from home (Elliott 2020). In the same year, the Welsh Government demonstrated a desire to preserve the increased prevalence of remote working spurred by the pandemic, aiming to have 30% of Welsh workers working from or near home (Welsh Government 2020a, b). This target aligns with estimates for global workforce remote working levels post-pandemic (Global Workplace Analytics 2020), and would result in Wales mirroring a handful of European Union countries’ pre-pandemic levels of remote working -a surplus of 30% of those in work teleworked at least sometimes in Sweden, Luxembourg, Finland and the Netherlands prior to the COVID-19 pandemic in 2019 (European Commission Joint Research Centre 2020).

While home working has increased, the availability of roles for which remote working is possible is largely dependent on the economic make-up of individual countries, and the distribution of roles across sectors. Resultantly, some individuals are well-positioned to benefit from a transition into home working, while others may be left disadvantaged. Higher-paid roles are more likely to allow for home-working, while workers within certain sectors are less likely to be offered the opportunity to work remotely e.g. those working within hospitality or retail (European Commission Joint Research Centre 2020).The COVID-19 pandemic has also highlighted how working from home is less accessible for key workers (Dyakova et al. 2021), those in precarious or low paid work (Dingel and Neiman 2020; Williams et al. 2020), and the digitally excluded (e.g. Bhandari 2020; Yates 2020). Between 2018 and 2019, 11% of Welsh adults could be categorized as digitally excluded, with those within households in the most deprived areas being less likely to have access to the internet than those in the least deprived areas (83% compared to 92% (Welsh Government 2019)). A transition into home-working has potential to both ameliorate and exacerbate inequalities for certain population groups. Establishing the extent to which different sub-groups are able to work from home, how home working impacted individuals’ health during the pandemic, and people’s preferences for future home-working will provide the insights needed to ensure that any policy-level changes that promote the continuation of home working protect against the potential damage to health and widening inequalities.

In this paper we use Welsh data to assess:

1. How does the ability to work from home differ across population sub-groups?
2. For individuals and sub-groups that are able to work from home, how does it impact their self-reported health?
3. How do the preferences for time spent working from home in future compare across population sub-groups?

## Methods

### Study design

A nationally-representative longitudinal household survey was undertaken across Wales (Public Health Wales’ *COVID-19, Employment and Health in Wales* study) with a paper-to-web push approach. The Health Research Authority approved the study (IRAS: 282223). Data was collected at two time-points. T1 data collection occurred in May-June 2020, and those that consented to participation in the follow-up at T2 were contacted again between December 2020 and January 2021.

### Study population and recruitment

All working age adults aged between 16-64 years resident in Wales, in current employment as of February 2020, were eligible, with those in full-time education or unemployed being excluded. To obtain a sample that was representative of the Welsh population, a stratified random probability sampling framework by age, gender and deprivation quintile was adopted. Respondents were informed that their participation was voluntary and that their responses would be confidential. Reminder letters were sent 10 days following original invitation. For each household, the eligible adult with the next birthday was asked to participate. A total of 1,382 adults responded at T1 (6.9% response rate), with 1,019 being from within the main sample (7.0% response rate), and 273 from the booster sample (5.5% response rate). Full details of the initial recruitment and sampling strategy are discussed elsewhere (Gray et al, 2021). Of the 1,382 adults who responded to the initial survey at T1, 1,084 individuals gave permission to be contacted for a follow up study. The follow-up data collection phase was from November 2020 to January 2021. If a valid email address was provided (N=925), individuals were emailed an invitation to take part with two further email remainders to encourage participation. If a valid email address was not provided (N=159), individuals were sent a postal invitation and one reminder invitation. In total, 626 individuals completed the follow-up online questionnaire (58% response rate). Our research explores questions only asked at T2, and therefore uses this sub-sample. Nine responses were excluded as identification codes were inputted incorrectly, leaving a sample of 615 (98.2%).

### Questionnaire measures

At T2, respondents were asked about their ability to work from home (‘able’ / ‘unable’ / ‘not sure’), the impact (‘better’, ‘no change’, ‘worse’) that home-working had on various aspects of their health (‘feelings of loneliness’, ‘mental well-being’, ‘smoking’, ‘eating well’, ‘drinking alcohol’, ‘exercise’, ‘work-life balance’), and their preference for the future (‘all working days home-working’, ‘half or more’, ‘less than half’, ‘no home-working’, ‘not sure’). Full details of these questionnaire measures are available in Supplementary Materials 1 (SM1).

To explore how the above-mentioned differed across population sub-groups, measurements from questions relating to socio-economic status, health and employment/income were also taken. Explanatory variables included age group, gender, deprivation quintile (assigned using the Welsh Index of Multiple Deprivation (Welsh Government 2021a) and residential postcode data), individual self-reported general health and presence of limiting pre-existing conditions (using validated questions from the National Survey for Wales (Welsh Government 2021b)), and mental well-being (using the short version of the Warwick Edinburgh Mental Well-being Scale (see Stewart-Brown et al. 2011) and using 1 SD below the mean as our cut-off score for low mental well-being). Explanatory variables relating to employment and income were also included, these were employment contract type (permanent, fixed term, atypical, self-employed/freelance), furlough status, wage precariousness (computed across three variables (see SM1) based on the Employment Precariousness Scale (Vives et al. 2015)) and job skill level (calculated using the Standard Occupational Classification for the UK (Office for National Statistics 2020)).

### Statistical approach

Statistical analysis was undertaken in IBM SPSS Statistics (Version 24) and R (Version 1.4.1103). Chi-square (χ^2^) and Fisher’s Exact tests were used to explore associations across socio-economic groups, employment and income, and health status. Multivariate binary and multinomial logistic regressions (adjusting for socio-economic factors, employment and income and self-reported health) were used to identify independent predictors of the ability to work from home, its health impacts, and preferences for future home-working.

## Results

### Sample characteristics

Respondents predominantly identified as women (63.7%). Those aged between 40 and 59 years of age were over-represented (40-49 24.6%; 50-59 32.7%), however there was representation across all other working ages. Responses were well distributed across deprivation quintiles (see SM2 for full sample characteristics).

### Research Question 1. How does the ability to work from home differ across population sub-groups?

Respondents who were unsure of whether they were able to work from home were excluded from the analysis, leaving a total of 580 respondents. Of these individuals, 51.6% reported that it was possible for them to work from home in their main job. The ability to work from home differed by socioeconomic factors, employment and income, and health (Table 1).

**Table 1.** Percentage of respondents reporting being able to work from home between November 2020 and January 2021, compared across socio-economic factors, employment and income, and health.

| Factors | N | % of which reporting being<br>able to<br>work from home | Significance<sup>(1)</sup> |
| --- | --- | --- | --- |
| *Gender* |  |  |  |
| Men | 201 | 47.8 | *p* = .19 |
| Women | 376 | 53.5 |  |
| Missing | 38 |  |  |
| *Age group* |  |  |  |
| 18-29 | 42 | 45.2 | *p* = .92 |
| 30-39 | 109 | 52.3 |  |
| 40-49 | 147 | 52.4 |  |
| 50-59 | 184 | 53.3 |  |
| 60-64 | 87 | 50.6 |  |
| Missing | 46 |  |  |
| *Deprivation quintile* |  |  |  |
| 1 | 107 | 43.0 | *p* = .003 |
| 2 | 140 | 42.1 |  |
| 3 | 92 | 52.2 |  |
| 4 | 104 | 60.6 |  |
| 5 | 137 | 60.6 |  |
| Missing | 35 |  |  |
| *Living arrangements* |  |  |  |
| Live alone | 111 | 44.1 | *p* = .09 |
| Not alone | 466 | 53.2 |  |
| Missing | 38 |  |  |
| *Children in household* |  |  |  |
| Children | 206 | 57.3 | *p* = .04 |
| No children | 374 | 48.4 |  |
| Missing | 35 |  |  |
| *Contract type* |  |  |  |
| Permanent | 464 | 53.2 | *p* = .001 |
| Fixed term | 33 | 60.6 |  |
| Atypical | 24 | 12.5 |  |
| Self-employed / Freelance | 58 | 48.3 |  |
| Missing | 36 |  |  |
| *Wage precarity*<sup>(2)</sup> |  |  |  |
| Low | 150 | 62.0 | *p* < .0001 |
| Moderate | 196 | 54.6 |  |
| High | 148 | 29.7 |  |
| Missing | 121 |  |  |
| *General health* |  |  |  |
| Good | 430 | 53.5 | *p* = .08 |
| Not good | 148 | 45.3 |  |
| Missing | 37 |  |  |
| *Mental well-being* |  |  |  |
| Low | 74 | 45.9 | *p* = .32 |
| Average | 502 | 52.2 |  |
| Missing | 39 |  |  |
| *Limiting pre-existing condition* |  |  |  |
| Yes | 124 | 43.5 | *p* = .03 |
| No | 428 | 54.7 |  |
| Missing | 63 |  |  |

*Note.* (1) *p* values calculated with Chi-square ( $\chi^2$ ) tests (2) Wage precarity calculated across three variables with between 4.5 and 8.5% of responses meeting criteria (see SM1), accounting for high level of missingness for composite variable.

#### Socio-economic factors and living arrangements

When controlling for all other factors within the model, women were nearly two times more likely to report being able to work from home than men (aOR 1.85 [95% CI 1.11-3.08]; see SM3 for full model outputs). As shown in Table 1, significant associations were found between the ability to work from home and deprivation (*p*=.003), as well as whether there were children in the household (p=.04).

#### Employment and Income

Less than 15% of those in atypical employment were able to work from home, with these individuals being less likely to be able to work from home than those in permanent employment (aOR 0.11 [95% CI 0.01-0.88]). In contrast, approximately 48% of the self-employed/freelancers could work from home, while 53.2% of those in permanent employment and 60.6% of those in fixed term employment could (*p=*.001). Those with high wage precarity were also less likely to be able to work from home than their counterparts with low wage precarity (aOR 0.29 [95% CI 0.15-0.55]).

#### Health status

Individuals who reported having a limiting pre-existing condition were less likely to be able to work from home than their healthier counterparts (43.5% compared to 54.7%, p=.03), however this effect was not significant when controlling for all other factors in the model (see SM3).

### Research Question 2: For individuals and sub-groups that are able to work from home, how does it impact their self-reported health?

Respondents who could work from home (N = 299) were asked to self-report how they thought working from home affected their health and well-being, including feelings of loneliness, mental well-being, smoking, diet, exercise, alcohol consumption, and work-life balance.

As shown in Figure 1, respondents who could work from home were more likely to report that it worsened their feelings of loneliness and mental well-being than to say it improved it, with over 40% indicating deteriorations for both aspects of health and well-being (and only 4.4% and 16.8% respectively reporting improvements). The same pattern was seen for alcohol consumption, with 23.6% reporting a deterioration, and only 5.5% reporting an improvement, and to a lesser extent, smoking (with 2.9% reporting an improvement and 7.4% reporting a deterioration). However, these patterns did differ across population sub-groups (see SM4 for proportions of individuals within each sub-group reporting each health outcome, and independent predictors of health outcomes as determined within multivariate logistic regression models).

**Figure 1.**
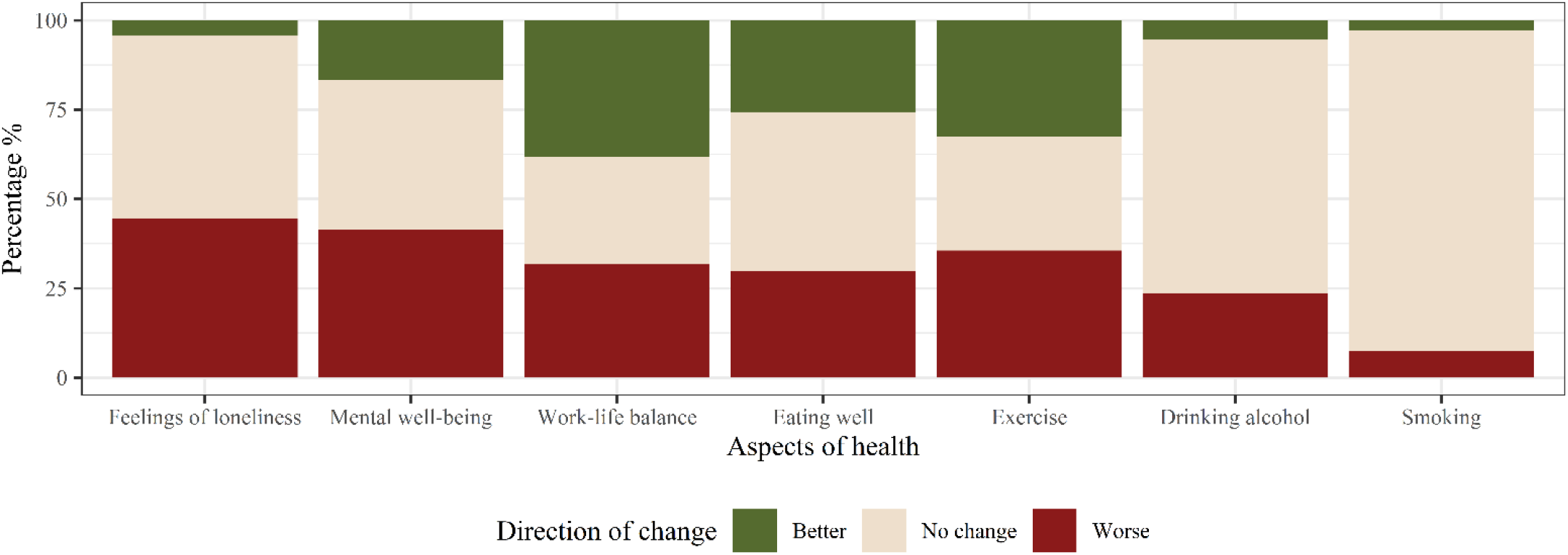
Percentage reporting improvements, no change or deteriorations in various aspects of their health and well-being as a result of home-working.

#### Socio-economic factors and living arrangements

Respondents under 50 years old were more likely to report a worsening in feelings of loneliness and their mental well-being as a result of working from home. When controlling for all other factors, it was found that those in their 30s were more than 3 times more likely than those in their 40s to report a worsening in their feelings of loneliness as a result of home-working (aOR 3.32 [95% CI 1.24-8.88]). Those in their 50s were significantly less likely to report deteriorations in their mental well-being than those in their 40s (aOR 0.30 [95% CI 0.11-0.82]), again corroborating the idea that younger individuals were more likely to see the negative impacts of home-working on their mental well-being.

Younger respondents were also more likely than older respondents to report that their diet worsened as a result of home-working, with those in their 30s being nearly five times more likely to report deteriorations in their diet than those in their 40s (aOR 4.65 [95% CI 1.44-14.44]). Similarly, when controlling for all other factors, individuals in their 30s were 6 times more likely than those in their 40s to report that working from home had a detrimental impact on their levels of physical activity (aOR 6.10 [95% CI 1.76-21.15]).

#### Employment and Income

Those in fixed term employment were less likely than those in permanent employment to report a deterioration in their sense of loneliness (aOR 0.10 [95% CI 0.02-0.61]). Those in permanent employment were most likely to report deteriorations in their mental well-being as a result of working from home (43.9%), while less than a third of all other groups reported such a worsening in their mental well-being (p=.01).

#### Health status

The multivariate model indicated that those in poorer health were more than 7 times more likely to report a deterioration in their diet (aOR 7.24 [95% CI 2.33-22.49]), over 5 times more likely to report a deterioration in their levels of physical activity (aOR 5.26 [95% CI 1.72-16.12]), nearly 8 times more likely to report deteriorations in their smoking habits (aOR 7.94 [95% CI 1.03-61.43]), and nearly 3 times more likely to report a worsening of levels of alcohol consumption (aOR 2.73 [95% CI 1.05-7.10]) when compared with their healthier counterparts.

Those with low mental well-being were more likely to report a deterioration in their feelings of loneliness as a result of home working (70.6% compared to 41.8%). Individuals with low mental well-being who worked from home were significantly more likely to report a deterioration in their sense of loneliness than their counterparts with average mental well-being (aOR 18.98 [95% CI 3.53-102.07]), with 70.6% of those with low mental well-being reporting becoming lonelier, and only 41.8% of their healthier counterparts reporting the same. Those with low well-being were also more likely to report a deterioration in their well-being (61.8% compared to 38.7%), with the model indicating that they were more than 4 times more likely to report deteriorations as a result of home-working (aOR 4.44 [95% CI 1.25-15.79]).

Associations suggested that those with limiting pre-existing conditions were more likely to report a change in their mental well-being as a result of home working (75.9% compared to 52.8%), with over half reporting a deterioration in their mental well-being (53.7% compared to 37.8%), and over a fifth reporting improvements (22.2% compared to 15%; p=.01). These associations were not found in the model when controlling for other factors. However, the model indicated that individuals with limiting pre-existing conditions were significantly less likely to report a worsening in their smoking habits, compared to their counterparts without such conditions (aOR 0.08 [95% CI 0.01-0.90]).

### Research Question 3: How do the preferences for time spent working from home in future compare across population sub-groups?

More than half the sample wanted to spend some of their working week working from home (53.2%), however only 13.2% wished to work from home all the time. A quarter of respondents indicated a desire to spend more than half their working week (but not the whole week) working from home in the future (25.9%), while 14.1% wished to spend less than half (but some of) the week working from home. A similar proportion indicated their desire to avoid working from home entirely (16.1%). There was also a degree of uncertainty, with over a fifth not sure of their preferences for future home working (22%). These proportions differed across the socio-economic, employment & income and health factors of interest (see SM5 for associations and multivariate multinomial regression model).

#### Socio-economic factors and living arrangements

Women were nearly three times more likely than men to want to work from home all the time instead of half the time (aOR 2.63 [95% CI 1.06-6.50]). In contrast, those in their 30s were less likely to report wanting to work from home all the time than those in their 40s (aOR 0.29 [95% CI 0.09-0.96]), and individuals who lived alone were more than two times more likely to not want to work from home at all (aOR 2.40 [95% CI 1.05-5.50]).

Individuals living within the second least deprived areas were less likely to report a preferences for no home working at all than those living in the least deprived areas (aOR 0.27 [95% CI 0.08-0.89]). Although not significant when controlling for other factors in the model, individuals living in the most deprived areas were less likely to report wanting to work from home all the time, more likely to report that they wanted to avoid working from home entirely, and more likely to show some uncertainty about future home working (p=.03).

#### Employment and Income

The self-employed/freelancers were nearly 7 times more likely than those in permanent employment to want to work from home all the time instead of half the time or more (aOR 6.98 [95% CI 1.98-24.59]). Working from home at least half of the working week was the preferred option for those in permanent or fixed term employment. Half of those in atypical employment were not sure of their preferences, and this group was the least likely to report a preference for full-time home working.

Those who were placed on furlough showed a greater level of uncertainty about their preference for future home-working, with 37.1% reporting they were not sure, compared to 20.9% for those not furloughed (p=.01) – this uncertainty was echoed in the model, with furloughed individuals being 3 times more likely to state they were uncertain of their preferences (aOR 3.09 [95% CI 1.40-6.82]).

Those with high levels of wage precarity demonstrated the greatest level of uncertainty about their preferences for future home working (39.3% compared to 22.2% for those with moderate wage precarity, and 16.6% for those with low levels), being more than four times more likely to state they were uncertain of their preference than those with low wage precarity (aOR 4.32 [95% CI 1.69-11.05]).

## Discussion

Our findings demonstrate that more than half of our Welsh sample are able to, and showed a preference for working from home to some capacity. These findings align with previous work with an UK-wide sample, indicating that nearly 90% of those that worked from home during the pandemic wished to continue doing so to some extent, with nearly half wishing to work from home either often or all the time (Felstead and Reuschke 2020). Within our sample, over a third wished to continue to work from home either full time or for half the working week or more. These findings could be seen to forecast that initiatives that seek to drive towards greater levels of home-working in future, such as the Welsh Government’s intention to have 30% of the Welsh workforce working remotely, are set to be well-received by workers (Welsh Government 2020a, b). For the majority, working from home is a viable and preferable option. Our study does however have three key implications for the development of policy promoting and supporting home working, specifically around addressing potential inequalities in home working, identifying and supporting the potential health impacts of home working, and flexing working arrangements and workplaces.

### Addressing inequalities in home working

Although these findings initially appear favourable for future initiatives, the ability to work from home is not evenly distributed across society. Within the context of the COVID-19 pandemic, the inaccessibility of home-working translates to increased exposure to the virus if remaining in-work, or financial insecurity if the work cannot continue e.g. being placed on furlough or becoming unemployed (Dyakova et al. 2021; Gray et al. 2021). Our findings corroborate those of others, showing that those living in the most deprived areas (who are also most likely to be digitally excluded (Welsh Government 2019)), those in atypical employment and those with high wage precarity are less likely to be able to work from home (Dingel and Neiman 2020; Williams et al. 2020). Although not significant when controlling for other factors in the model, a significant association within this sample showed that those with limiting pre-existing conditions were less likely to be able to work from home. Provisions that can be put in place at the workplace to support employee needs may not be as easily adapted in the home, with research suggesting that equipment used during working from home is less ergonomically suitable, with this affecting work performance for individuals with disabilities (Ralph et al. 2020; Guler et al. 2021). Wage precarity and atypical working arrangements are associated with increased risks of experiencing ill-health and financial insecurity (Benach et al. 2014). Individuals with poorer health or limiting conditions are already at a disadvantage in obtaining and retaining work due to the challenges that their symptoms and their treatment needs present (Khan et al. 2009; Mack and Paylor 2016; van Egmond et al. 2016; Brannigan et al. 2017; Nexo et al. 2017; Booth et al. 2018; Hanson et al. 2018; Paltrinieri et al. 2018; Murray et al. 2019).

Taken together, a more permanent transition towards home-working may cause further insecurity and exclusions for sub-groups that are experiencing greater financial insecurity and ill-health, who are at present less able to participate in this change. Supporting individuals with specific work-related needs to address them within the home-working environment might help. Further drives to promote working from home that do not account for discrepancies in the accessibility of home-working may widen existing inequalities. In the context of COVID-19 or another pandemic response, widening the accessibility of home-working for these highlighted groups where possible will help protect their health. Where roles cannot be performed at home (e.g. key workers, retail), the increased risk of infection that these sub-groups face as a result of their inability to work from home should be considered when designing support packages at a population level.

### Identifying and supporting the health impacts of home-working

A second concern for the wider adoption of home-working is its health impacts. Although home-working can protect individuals from exposure to communicable diseases as described above, working from home can present its own health challenges. Of those that were able to work from home, more than 40% reported that their mental well-being and their sense of loneliness deteriorated as a result of home-working. These findings align with those of the Understanding Society Covid-19 Study, which showed that those that worked from home full-time during the UK’s first national lockdown reported significant deteriorations in their well-being, including their enjoyment of normal activities, experiences of strain, their ability to concentrate and experiencing unhappiness/depression (Felstead and Reuschke 2020).

Of note, Felstead and Reuschke’s work showed that those that worked from home part-time had significantly better outcomes, and those home-working full-time became less severely affected by June 2020 (compared to April and May), with the authors suggesting that individuals either became accustomed to home-working or were able to return to work if it had severely affected their mental well-being. With our Welsh data gathered between November 2020 and January 2021, it would appear that the detrimental impacts of home-working persisted later into the year, with this perhaps indicating a combined effect of home-working, the winter months and government restrictions. With those adopting a hybrid home-working model being less affected (as reported by Felstead and Reuschke (2020), pursuing the Welsh public’s preference for a hybrid model (with 40% of our sample wanting a hybrid approach) would contribute to protecting against the negative impacts on well-being that full-time home-working might produce. Caution must be taken to avoid the negative health outcomes that our sample reported experiencing, particularly for their mental well-being. Our findings also suggest that younger individuals (under 50 years of age) and those with low mental-wellbeing are more likely to report experiencing these detriments to their mental well-being and sense of loneliness when working from home. Processes that allow for regular review of full-time home-workers’ health may prove beneficial in boosting the effectiveness of the adoption of home-working on a longer term basis, as would providing targeted support for groups that are more likely to report feelings of isolation or see their mental well-being deteriorate (e.g. peer support groups, access to work networks, advice and guidance).

Alcohol consumption and smoking habits were also more likely to worsen than improve for those working from home. Similar patterns have been found by others for both alcohol consumption (e.g. 30% of a 2,777 self-selected UK sample reported drinking more frequently during lockdown (Oldham et al. 2021) and harmful alcohol use increased for those working from home under lockdown in the US between April and September 2020 (Killgore et al. 2021)) and smoking (e.g. 28% of an US sample of 291 tobacco users reported increasing their cigarette use during the pandemic, reporting increased time at home as one of their reasons (Yingst et al. 2021)). As suggested by Killgore et al. (2021), the increased freedoms and privacy that home-working can offer opens the door for engaging in behaviours that would otherwise be reserved for outside of working hours or working environments. While hybrid approaches would reduce these impacts, employees may require additional support in maintaining workplace standards within the home environment. Home-worker health reviews could signpost to resources for support in curbing unhealthy habits.

Home-working appeared to detrimentally impact many sub-groups’ diets and levels of physical activity – a pattern already highlighted as a risk of home-working in previous work (Bevan et al. 2020). In our sample, those in their 30s and those with poorer general health were more likely to report these effects. Our findings highlight an important issue, with the data indicating that home-working introduced additional health challenges to those with poorer general and mental health. Individuals with poorer general health were seven times more likely to report a worsened diet as a result of home-working, over five times more likely to report a deterioration in their levels of physical activity, and reported a worsening in their engagement in health-harming behaviours (alcohol consumption and smoking). In addition to being more likely to experience deteriorations in their sense of loneliness and their mental well-being, individuals with low mental-wellbeing also saw their diets suffer. The disruptive shift to home-working may have interrupted many individuals’ pre-existing habits and routines. While other research has suggested that the negative impacts of home-working on diet and physical activity may subside as individuals get accustomed to these work-related changes (e.g. Rogers et al. 2021), efforts should be made to support individuals in establishing healthier habits while working from home, particularly those individuals that may be balancing efforts to maintain such habits while attending to their other health needs. The disproportionate self-reported negative health impacts of home-working on those with poorer health are of particular concern when considering the fact that working from home is associated with increased sickness presenteeism (Karanikas and Cauchi 2020; Steidelmüller et al. 2020), which holds the potential to worsen existing health problems.

### Flexing working arrangements and workplaces

For the last of the three themes covered in this research, our findings highlight how a wider adoption of home-working should account for how different sub-groups have different preferences for the time they spend working from home. While efforts should be made to increase the accessibility of home-working for men, who were significantly less likely to be able to work from home than women, women were significantly more likely to want to work from home throughout the working week. The flexibility that home-working can offer is well-suited for a population group that are more likely to carry the burden of caring responsibilities. In contrast, those in their 30s, those that live alone and the most deprived were significantly more likely to want to spend some or all of their time working from an office/base instead of at home. For the former two, working from a base would presumably allow for greater opportunities for socialisation. Research has suggested that those living alone are more likely to experience mental illness (McManus et al. 2014). Work-related situational constraints that affect the extent to which individuals spend time within physical proximity to colleagues, or the frequency of their interactions, can affect employees’ sense of isolation (Perlman and Peplau 1981).

With home-working associated with increased loneliness and detriments to well-being within our sample, initiatives that promote further remote or home working could protect against isolation through adopting hybrid approaches, supporting individuals in accessing remote working hubs or through ensuring that employers provide home-workers with support in maintaining contact with colleagues. As discussed, younger individuals and those with low mental-wellbeing were more likely to report deteriorations in their mental well-being and their sense of loneliness as a result of home-working. These groups in particular might benefit from having the option to work in an office/base, in a remote working hub, or with a hybrid model combining working on and off site.

### Strengths and limitations

Our findings relating to home-working’s health impacts are limited due to the cross-sectional nature of our study, and the fact that these were self-reported measures taken during the COVID-19 pandemic. For many in our sample, their experiences of home-working came as a direct consequence of the pandemic, which in itself contributed to feelings of isolation, presented increased burdens on mental well-being, and impacted people’s engagements in many health-related behaviours. Home-working coupled with these may have produced the effects seen here. While this could signify that our findings are best applied in informing future pandemic responses, they do provide a representative overview of the groups that might not be able to participate in the transition towards home-working, and the groups that are most likely to experience its less beneficial health outcomes. These insights can contribute to ensuring that future policies and practice that promote working from home (be that for future pandemic responses or as part of efforts to tackle climate change) adopt preventative measures which contribute to protecting these groups from experiencing increased isolation, seeing their mental well-being decline, increasing health-harming behaviours and decreasing engagement in healthy behaviours. Generating knowledge which allows for identifying those that are at risk of the negative health impacts of home-working can help inform the development of targeted support for these sub-groups going forward.

## Conclusion

Individuals with high wage precarity, those on atypical contracts and those living in the most deprived areas were less likely to be able to work from home. The ability to work from home is not equally distributed within Wales, with those facing greatest insecurity being more likely to lose out. While home-working offers many benefits, many have seen it deteriorate their mental well-being and sense of loneliness, with this being particularly true for younger individuals and those with low mental well-being. Less healthy consumption behaviours and more sedentary lifestyles were also results of home-working for younger groups and those in poorer health. With over 50% of our sample able to work from home and indicating a preferences towards doing so to some capacity, the public health challenge that these negative health impacts present is clear. The push for a more permanent transition to working from home should account for these inequities in access, and protect against the potential detriments to health.

## Data Availability

All data produced in the present study are available upon reasonable request to the authors

## References

Arshed N, Meo MS, Farooq F (2020) Empirical assessment of government policies and flattening of the COVID19 curve. Journal of Public Affairs 20:e2333. https://doi.org/10.1002/pa.2333

Bachelet M, Kalkuhl M, Koch N (2021) What If Working from Home Will Stick? Distributional and Climate Impacts for Germany. SSRN. https://doi.org/10.2139/ssrn.3908857

Beatty JE, Joffe R (2006) An Overlooked Dimension Of Diversity: The Career Effects of Chronic Illness. Organizational Dynamics 35:182–195. https://doi.org/10.1016/j.orgdyn.2006.03.006

Benach J, Vives A, Amable M, et al (2014) Precarious employment: Understanding an emerging social determinant of health. Annual Review of Public Health 35:229–253. https://doi.org/10.1146/annurev-publhealth-032013-182500

Beno M (2021) Face-to-display working: Decarbonisation potential of not commuting to work before Covid-19 and during and after lockdowns. Academic Journal of Interdisciplinary Studies 10:17–24. https://doi.org/10.36941/AJIS-2021-0060

Bevan S, Mason B, Bajorek Z (2020) Homeworker Wellbeing Survey. https://www.employment-studies.co.uk/sites/default/files/resources/summarypdfs/IES%20Homeworker%20Wellbeing%20Survey%20-%20Interim%20Findings.pdf. Accessed 11 Nov 2021

Bhandari V (2020) Improving internet connectivity during Covid-19. SSRN. https://doi.org/10.2139/ssrn.3688762

Booth S, Price E, Walker E (2018) Fluctuation, invisibility, fatigue - the barriers to maintaining employment with systemic lupus erythematosus: results of an online survey. Lupus 27:2284–2291. https://doi.org/10.1177%2F0961203318808593

Brannigan C, Galvin R, Walsh ME, et al (2017) Barriers and facilitators associated with return to work after stroke: a qualitative meta-synthesis. Disability and Rehabilitation 39:211–222. https://doi.org/10.3109/09638288.2016.1141242

Dingel JI, Neiman B (2020) How Many Jobs Can be Done at Home? Journal of Public Economics 189:104235. https://doi.org/10.1016/j.jpubeco.2020.104235

Dyakova M, Couzens L, Allen J, et al (2021) Placing health equity at the heart of the COVID-19 sustainable response and recovery: Building prosperous lives for all in Wales. https://phw.nhs.wales/news/placing-health-equity-at-the-heart-of-coronavirus-recovery-for-building-a-sustainable-future-for-wales/placing-health-equity-at-the-heart-of-the-covid-19-sustainable-response-and-recovery-building-prosperous-lives-for-all-in-wales. Accessed 4 Jan 2022

Elliott D (2020) Germany drafting law to give people the legal right to work from home. In: World Economic Forum. https://www.weforum.org/agenda/2020/10/germany-is-set-to-make-home-working-a-legal-right/. Accessed 2 Dec 2021

Eurofound (2020) Living, working and COVID-19. In: Publications Office of the European Union. https://www.eurofound.europa.eu/sites/default/files/ef_publication/field_ef_document/ef20059en.pdf. Accessed 5 Jan 2022

European Commission Joint Research Centre (2020) Telework in the EU before and after the COVID-19: where we were, where we head to. https://ec.europa.eu/jrc/sites/default/files/jrc120945_policy_brief_-_covid_and_telework_final.pdf. Accessed 2 Dec 2021

Felstead A, Reuschke D (2020) Homeworking in the UK: before and during the 2020 lockdown. https://wiserd.ac.uk/publications/homeworking-uk-and-during-2020-lockdown. Accessed 11 Nov 2021

Global Workplace Analytics (2020) Work-At-Home After Covid-19 - Our Forecast. https://globalworkplaceanalytics.com/work-at-home-after-covid-19-our-forecast. Accessed 2 Dec 2021

Gray BJ, Kyle RG, Song J, Davies AR (2021) Characteristics of those most vulnerable to employment changes during the COVID-19 pandemic: A nationally representative cross-sectional study in Wales. Journal of Epidemiology and Community Health 76:8–15. https://doi.org/10.1136/jech-2020-216030

Guler MA, Guler K, Guneser Gulec M, Ozdoglar E (2021) Working From Home During a Pandemic: Investigation of the Impact of COVID-19 on Employee Health and Productivity. Journal of Occupational and Environmental Medicine 63:731–741. https://doi.org/10.1097/JOM.0000000000002277

Hanson H, Hart RI, Thompson B, et al (2018) Experiences of employment among young people with juvenile idiopathic arthritis: a qualitative study. Disability and Rehabilitation 40:1921–1928. https://doi.org/10.1080/09638288.2017.1323018

Holland P, Collins AM (2018) “Whenever I can I push myself to go to work”: a qualitative study of experiences of sickness presenteeism among workers with rheumatoid arthritis. Disability and Rehabilitation 40:404–413. https://doi.org/10.1080/09638288.2016.1258436

Hook A, Court V, Sovacool BK, Sorrell S (2020) A systematic review of the energy and climate impacts of teleworking. Environmental Research Letters 15:093003. https://doi.org/10.1088/1748-9326/ab8a84

Karanikas N, Cauchi JP (2020) Literature review on parameters related to Work-From-Home (WFH) arrangements. https://eprints.qut.edu.au/205308/. Accessed 5 Jan 2022

Khan F, Ng L, Turner-Stokes L (2009) Effectiveness of vocational rehabilitation intervention on the return to work and employment of persons with multiple sclerosis. Cochrane Database of Systematic Reviews 1:CD007256. https://doi.org/10.1002%2F14651858.CD007256.pub2

Killgore WDS, Cloonan SA, Taylor EC, et al (2021) Alcohol dependence during COVID-19 lockdowns. Psychiatry Research 296:113676. https://doi.org/10.1016/j.psychres.2020.113676.

Kramer A, Kramer KZ (2020) The potential impact of the Covid-19 pandemic on occupational status, work from home, and occupational mobility. Journal of Vocational Behavior 119:103442. https://doi.org/10.1016/j.jvb.2020.103442

Mack H, Paylor I (2016) Employment Experiences of Those Living with and Being Treated for Hepatitis C: Seeking Reasonable Adjustments and the Role of Disability Legislation. Social Policy and Society 15:555–570. https://doi.org/10.1017/S1474746415000378

McManus S, Bebbington P, Jenkins R, Brugha T (2014) Mental health and wellbeing in England: Adult Psychiatric Morbidity Survey. https://openaccess.city.ac.uk/id/eprint/23646/1/. Accessed 16 Dec 2021

Miles D, Stedman M, Heald AH (2021) “Stay at Home, Protect the National Health Service, Save Lives”: a cost benefit analysis of the lockdown in the United Kingdom. International Journal of Clinical Practice 75:e13674. https://doi.org/10.1111/ijcp.13674

Murray PD, Brodermann MH, Gralla J, Wiseman AC (2019) Academic achievement in employment in young adults with end stage kidney disease. Journal of Renal Care 45:29–40. https://doi.org/10.1111/jorc.12261

Nexo M, Cleal B, Hagelund L, et al (2017) Willingness to pay for flexible working conditions of people with type 2 diabetes: Discrete choice experiments. BMC Public Health 17:938. https://doi.org/10.1186/s12889-017-4903-6

Office for National Statistics (2020) Standard Occupational Classification - SOC 2020. https://www.ons.gov.uk/methodology/classificationsandstandards/standard. Accessed 13 Oct 2021

Oldham M, Garnett C, Brown J, et al (2021) Characterising the patterns of and factors associated with increased alcohol consumption since COVID-19 in a UK sample. Drug and Alcohol Review 40:890–899. https://doi.org/10.1111/dar.13256

Paltrinieri S, Fugazzaro S, Bertozzi L, et al (2018) Return to work in European Cancer survivors: a systematic review. Supportive Care in Cancer 26:2983–2994. https://doi.org/10.1007/s00520-018-4270-6

Perlman D, Peplau LA (1981) Toward a Social Psychology of Loneliness. In: Duck S, Gilmour R (eds) Personal Relationships in Disorder. Academic Press, London, pp 31–56

Ralph P, Baltes S, Adisaputri G, et al (2020) Pandemic programming: How COVID-19 affects software developers and how their organizations can help. Empirical Software Engineering 25:4927–4961. https://doi.org/10.1007/s10664-020-09875-y

Rogers AM, Lauren BN, Baidal JAW, et al (2021) Persistent effects of the COVID-19 pandemic on diet, exercise, risk for food insecurity, and quality of life: A longitudinal study among U.S. adults. Appetite 167:105639. https://doi.org/10.1016/j.appet.2021.105639

Rubin O, Nikolaeva A, Nello-Deakin S, te Brömmelstroet M (2020) What can we learn from the COVID-19 pandemic about how people experience working from home and commuting? In: Centre for Urban Studies, University of Amsterdam. https://www.researchgate.net/profile/Ori-Rubin/publication/341233510_What_can_we_learn_from_the_COVID-19_pandemic_about_how_people_experience_working_from_home_and_commuting/links/5eb5209b4585152169be9123/What-can-we-learn-from-the-COVID-19-pandemic-about-how-people-experience-working-from-home-and-commuting.pdf. Accessed 16 Dec 2021

Steidelmüller C, Meyer SC, Muller G (2020) Home-based telework and presenteeism across Europe. Journal of Occupational and Environmental Medicine 62:998–1005. https://doi.org/10.1097/JOM.0000000000001992

Stewart-Brown S, Platt S, Tennant A, et al (2011) The Warwick-Edinburgh Mental Well-being Scale (WEMWBS): a valid and reliable tool for measuring mental well-being in diverse populations and projects. Journal of Epidemiology and Community Health 65:A1–A40. https://doi.org/10.1136/jech.2011.143586.86

UK Government (2021) Government response: Health is everyone’s business. https://www.gov.uk/government/consultations/health-is-everyones-business-proposals-to-reduce-ill-health-related-job-loss/outcome/government-response-health-is-everyones-business. Accessed 14 Oct 2021

van Aerden K, Gadeyne S, Vanroelen C (2017) Is any job better than no job at all? Studying the relations between employment types, unemployment and subjective health in Belgium. Archives of Public Health 75:55. https://doi.org/10.1186/s13690-017-0225-5

van Egmond MP, Duijts SFA, Scholten APJ, et al (2016) Offering a tailored return to work program to cancer survivors with job loss: A process evaluation. BMC Public Health 16:940. https://doi.org/10.1186/s12889-016-3592-x

Vives A, González F, Moncada S, et al (2015) Measuring precarious employment in times of crisis: the revised employment precariousness scale (EPRES) in Spain. Gaceta Sanitaria 29:379–382. https://doi.org/10.1016/j.gaceta.2015.06.008

Waddell G, Burton AK (2006) Is work good for your health and well-being? TSO, London

Welsh Government (2020a) Embedding Remote Working. https://gov.wales/written-statement-embedding-remote-working. Accessed 14 Oct 2021

Welsh Government (2020b) Aim for 30% of the Welsh workforce to work remotely. https://gov.wales/aim-30-welsh-workforce-work-remotely#:~:text=Aim%20for%2030%20of%20the%20Welsh%20workforce%20to,home%2C%20including%20after%20the%20threat%20of%20Covid-19%20lessens. Accessed 11 Nov 2021

Welsh Government (2019) National Survey for Wales, 2018-2019 Internet use and digital skills. https://gov.wales/sites/default/files/statistics-and-research/2019-09/internet-use-and-digital-skills-national-survey-wales-april-2018-march-2019-207.pdf. Accessed 11 Nov 2021

Welsh Government (2021a) Welsh Index of Multiple Deprivation. https://gov.wales/welsh-index-multiple-deprivation. Accessed 13 Oct 2021

Welsh Government (2021b) National Survey for Wales. https://gov.wales/national-survey-wales. Accessed 13 Oct 2021

Williams S, Armitage C, Tampe T, Dienes K (2020) Public perceptions and experiences of social distancing and social isolation during the COVID-19 pandemic: a UK-based focus group study. BMJ Open 10:e039334. https://doi.org/10.1136/bmjopen-2020-039334

Yates S (2020) COVID-19 and Digital Exclusion: Insights and Implications for the Liverpool City Region. https://livrepository.liverpool.ac.uk/3110809/1/Heseltine%20Institute%20Policy%20Briefing%20031.pdf. Accessed 16 Dec 2021

Yingst JM, Krebs NM, Bordner CR, et al (2021) Tobacco Use Changes and Perceived Health Risks among Current Tobacco Users during the COVID-19 Pandemic. International Journal of Environmental Research and Public Health 18:1795. https://doi.org/10.3390/ijerph18041795

