## Supplementary Material 1 for "Exploring the health impacts and inequalities of the new way of working: findings from a cross-sectional study"

### Supplementary Material 1: Questionnaire measures

Ability to work from home:

Is it possible in your main job role to work from home?

❑ Yes

❑ No

❑ Not sure

Preferences for future home-working:

Would you like the change in frequency of working from home to continue after COVID-19?

❑ I’d like to work from home on all of my working days

❑ I’d like to work from home on at least half of my working days

❑ I’d like to work from home on less than half of my working days

❑ I’d like to work from an office/base and not work from home

❑ Not sure

Health impacts:

How does working from home effect these aspects of your health and wellbeing?

Better	No change	Worse

Feeling of loneliness	❑	❑	❑

Mental wellbeing	❑	❑	❑

Smoking	❑	❑	❑
Eating well	❑	❑	❑

Drinking alcohol	❑	❑	❑

Exercise	❑	❑	❑

Work-life balance	❑	❑	❑

Wage precariousness computation:

Responses for three measures were used to compute wage precariousness. These included:

a. Thinking about your main job, what is your total personal income* from all sources?

❑ Less than £200 a week / less than £870 a month / less than £10,400 a year

❑ £200 to £399 a week / £870 to £1,729 a month / £10,400 to £20,799 a year

❑ £400 to £599 a week / £1,730 to £2,599 a month / £20,800 to £31,099 a year

❑ £600 to £799 a week / £2,600 to £3,459 a month / £31,100 to £41,499 a year

❑ £800 or more a week / £3,460 or more a month / £41,500 or more a year

❑ Don’t know

❑ Prefer not to say

*This is your own gross income – before any deductions like tax, national insurance, pension etc is taken off

To what extent does your income from your main job enable you to…

Always	Most of the time	Sometimes	Rarely	Never
b. cover your basic needs,

such as food, clothes,	❑	❑	❑	❑	❑

heating and housing costs?

c. cover unforeseen expenses,

e.g. urgent repair to a car,

replacement of household	❑	❑	❑	❑	❑

appliances etc?

Respondents who had omitted an answer for any of the three questions were excluded from the calculation (a: 8.5% missing; b: 4.9% missing; c: 5.5% missing; combined: 18.7% missing). Questions b and c were recoded onto a 0-4 scale (0 = always, 4 never). Scores for each of the three items were then divided by 12, summed, then multiplied by 4 to give a composite wage precariousness score. Scores below 1 indicate low wage precarity, scores between 1 and 1.99 indicated moderate wage precarity, and scores of 2 or above indicated high or very high wage precarity (i.e. higher financial insecurity).
