## Supplementary Material 2 for "Exploring the health impacts and inequalities of the new way of working: findings from a cross-sectional study"

### Supplementary Material 2: Sample characteristics

Table 1. Characteristics of the sample (including number and percentage of respondents for each factor level)

| **Factor** | **N** | **%** |
| --- | --- | --- |
| *Gender* |  |  |
| Man | 218 | 35.4 |
| Woman | 392 | 63.7 |
| Other | 2 | 0.3 |
| Missing | 3 | 0.5 |
| *Age group* |  |  |
| 18-29 | 43 | 7.0 |
| 30-39 | 112 | 18.2 |
| 40-49 | 151 | 24.6 |
| 50-59 | 201 | 32.7 |
| 60-64 | 96 | 15.6 |
| Missing | 12 | 2.0 |
| *Deprivation quintile* |  |  |
| 1 (Most deprived) | 114 | 18.5 |
| 2 | 150 | 24.4 |
| 3 | 95 | 15.4 |
| 4 | 113 | 18.4 |
| 5 (Least deprived) | 143 | 23.3 |
| Missing | 0 | 0 |
| *Living arrangements* |  |  |
| Live alone | 119 | 19.3 |
| Live with others | 492 | 80.0 |
| Missing | 4 | 0.7 |
| *Children in households* |  |  |
| Children | 211 | 34.3 |
| No children | 404 | 65.7 |
| Missing | 0 | 0 |
| *Contract type* |  |  |
| Permanent | 467 | 75.9 |
| Fixed term | 34 | 5.5 |
| Atypical | 25 | 4.1 |
| Self-employed / Freelance | 59 | 9.6 |
| Missing | 30 | 4.9 |
| *Furlough* |  |  |
| Yes | 116 | 18.9 |
| No | 478 | 77.7 |
| Missing | 21 | 3.4 |
| *Wage precarity* |  |  |
| Low | 151 | 24.6 |
| Moderate | 200 | 32.5 |
| High | 149 | 24.2 |
| Missing | 115 | 18.7 |
| *General health* |  |  |
| Good | 454 | 73.8 |
| Not good | 159 | 25.9 |
| Missing | 2 | 0.3 |
| *Mental well-being* |  |  |
| Low | 78 | 12.7 |
| Average | 528 | 85.9 |
| Missing | 9 | 1.5 |
| *Limiting pre-existing condition* |  |  |
| Yes | 132 | 21.5 |
| No | 454 | 73.8 |
| Missing | 29 | 4.7 |
