## Supplementary Material 3 for "Exploring the health impacts and inequalities of the new way of working: findings from a cross-sectional study"

### Supplementary Material 3: Multivariate logistic regression for ability to work from home

Table 1. Multivariate logistic regression model identifying independent predictors of ability to work from home.

|  | Ability to WFH (Valid N = 413) |
| --- | --- |
| *Gender* |  |
| Men | Reference |
| Women | **1.85 [1.11-3.08]**  **p = .02** |
| *Age group* |  |
| 18-29 Years | 0.82 [0.33-2.08]  p = .68 |
| 30-39 Years | 0.80 [0.41-1.55]  p = .51 |
| 40-49 Years | Reference |
| 50-59 Years | 1.19  [0.61-2.29] p = .61 |
| 60-64 Years | 1.79  [0.79-4.10]  p = .17 |
| *Deprivation quintile* |  |
| 1 Most deprived | 0.69  [0.34-1.40]  p = .30 |
| 2 | 0.82  [0.44-1.55]  p = .54 |
| 3 | 0.69  [0.34-1.41]  p = .31 |
| 4 | 0.99  [0.49-2.01]  p = .98 |
| 5 Least deprived | Reference |
| *Living arrangements* |  |
| Live alone | 0.83  [0.46-1.51] p = .55 |
| Live with others | Reference |
| *Children in household* |  |
| Children | 1.53  [0.86-2.73] p = .15 |
| No children | Reference |
| *Contract type* |  |
| Permanent | Reference |
| Fixed term | 1.47  [0.55-3.91] p = .44 |
| Atypical | **0.11 [0.01-0.88] p = .04** |
| Self-employed / Freelance | 1.27 [0.55-2.93] p = .57 |
| *Furlough* |  |
| Yes | **0.49**  **[0.27-0.90] p = .02** |
| No | Reference |
| Wage precarity |  |
| Low | Reference |
| Moderate | 0.76  [0.44-1.32] p = .33 |
| High | **0.29**  **[0.15-0.55] p < .001** |
| General health |  |
| Good | Reference |
| Not good | 1.19  [0.65-2.20]  p = .58 |
| Mental well-being |  |
| Low | 1.31  [0.61-2.82]  p = .49 |
| Average | Reference |
| Limiting pre-existing condition |  |
| Yes | 0.79  [0.44-1.45]  p = .45 |
| No | Reference |

*Note: Odds ratios adjusted for: gender, age, deprivation quintile, living arrangements, children in household, highest qualification level, contract type, furlough, wage precarity, job skill level, general health, mental well-being and limiting pre-existing conditions.*
