## Supplementary Material 4 for "Exploring the health impacts and inequalities of the new way of working: findings from a cross-sectional study"

### Supplementary Material 4: Health impacts of home-working

Table 1. Percentage of respondents that were able to work from home that indicated improvements (+), no change (0), or deteriorations (-) in various aspects of their health as a result of home-working.

| *Factors* | | *Health impacts of home-working* | | | | | | | | | | | | | | | | | | | | | | | | | | | | | | | | | | | | | | | |
| --- | --- | --- | --- | --- | --- | --- | --- | --- | --- | --- | --- | --- | --- | --- | --- | --- | --- | --- | --- | --- | --- | --- | --- | --- | --- | --- | --- | --- | --- | --- | --- | --- | --- | --- | --- | --- | --- | --- | --- | --- | --- |
|  | | *Feeling of loneliness* | | | | *Mental*  *well-being* | | | | | | | | | | *Smoking* | | | | *Eating well* | | | | | | | | | *Drinking alcohol* | | | | | *Exercise* | | | | | *Work-life balance* | | |
|  | | *+* | | *0* | *-* | *+* | | | | *0* | *-* | | | | | *+* | | *0* | *-* | *+* | | | | *0* | *-* | | | | *+* | | *0* | *-* | | *+* | | *0* | *-* | | *+* | *0* | *-* |
| *Gender* | | *N = 296 Missing = 3* | | | | *N = 296 Missing = 3* | | | | | | | | | | *N = 270 Missing = 29* | | | | *N = 294 Missing = 5* | | | | | | | | | *N = 290 Missing = 9* | | | | | *N = 296 Missing = 3* | | | | | *N = 296 Missing = 3* | | |
| Men  N = 96 | | 5.2% | | 56.3% | 38.5% | 16.7% | | | | 44.8% | 38.5% | | | | | 6.6% | | 87.9% | 5.5% | 26.0% | | | | 46.9% | 27.1% | | | | 6.3% | | 67.7% | 26.0% | | 34.4% | | 29.2% | 36.5% | | 44.8% | 27.1% | 28.1% |
| Women  N = 200 | | 3.5% | | 48.5% | 48.0% | 16.0% | | | | 41.0% | 43.0% | | | | | 1.1% | | 90.5% | 8.4% | 24.7% | | | | 43.9% | 31.3% | | | | 5.2% | | 72.2% | 22.7% | | 32.0% | | 33.0% | 35.0% | | 35.0% | 31.5% | 33.5% |
| p value | | *.26* | | | | *.76* | | | | | | | | | | ***.04*** | | | | *.76* | | | | | | | | | *.73* | | | | | *.80* | | | | | *.27* | | |
| *Age Group* | | *N = 294 Missing = 5* | | | | *N = 294 Missing = 5* | | | | | | | | | | *N = 268 Missing = 31* | | | | *N = 292 Missing = 7* | | | | | | | | | *N = 288 Missing = 11* | | | | | *N = 294 Missing = 5* | | | | | *N = 294 Missing = 5* | | |
| 18-29 Years  N = 19 | | 5.3% | | 31.6% | 63.2% | 21.1% | | | | 31.6% | 47.4% | | | | | 0 | | 94.7% | 5.3% | 36.8% | | | | 26.3% | 36.8% | | | | 5.3% | | 63.2% | 31.6% | | 26.3% | | 26.3% | 47.4% | | 31.6% | 31.6% | 36.8% |
| 30-39 Years  N = 57 | | 1.8% | | 42.1% | 56.1% | 19.3% | | | | 29.8% | 50.9% | | | | | 1.8% | | 85.7% | 12.5% | 26.3% | | | | 26.3% | 47.4% | | | | 8.8% | | 64.9% | 26.3% | | 29.8% | | 26.3% | 43.9% | | 35.1% | 22.8% | 42.1% |
| 40-49 Years  N = 77 | | 1.3% | | 48.1% | 50.6% | 14.3% | | | | 36.4% | 49.4% | | | | | 2.8% | | 91.5% | 5.6% | 27.3% | | | | 40.3% | 32.5% | | | | 6.8% | | 67.1% | 26.0% | | 41.6% | | 31.2% | 27.3% | | 41.6% | 29.9% | 28.6% |
| 50-59 Years  N = 98 | | 5.1% | | 61.2% | 33.7% | 12.2% | | | | 54.1% | 33.7% | | | | | 3.5% | | 88.4% | 8.1% | 20.8% | | | | 57.3% | 21.9% | | | | 4.1% | | 74.2% | 21.6% | | 27.6% | | 34.7% | 37.8% | | 39.8% | 27.6% | 32.7% |
| 60-64 Years  N = 43 | | 11.6% | | 53.5% | 34.9% | 25.6% | | | | 46.5% | 27.9% | | | | | 5.6% | | 91.7% | 2.8% | 27.9% | | | | 55.8% | 16.3% | | | | 2.4% | | 81.0% | 16.7% | | 32.6% | | 39.5% | 27.9% | | 37.2% | 44.2% | 18.6% |
| p value | | ***.01*** | | | | ***.04*** | | | | | | | | | | *.72* | | | | ***.003*** | | | | | | | | | *.69* | | | | | *.34* | | | | | *.31* | | |
| *Deprivation  Quintile* | | *N = 298 Missing = 1* | | | | *N = 298 Missing = 1* | | | | | | *N = 272 Missing = 27* | | | | | | | | *N = 296 Missing = 3* | | | | | | | | | | *N = 292 Missing = 7* | | | | *N = 298 Missing = 1* | | | | *N = 298 Missing = 1* | | | |
| 1 Most  N = 46 | | 2.2% | | 47.8% | 50.0% | 10.9% | | | | 47.8% | 41.3% | | | | | 7.1% | | 88.1% | 4.8% | 22.2% | | | | 33.3% | 44.4% | | | | 2.2% | | 75.6% | 22.2% | | 21.7% | | 28.3% | 50.0% | | 41.3% | 21.7% | 37.0% |
| 2  N = 59 | | 6.8% | | 52.5% | 40.7% | 16.9% | | | | 44.1% | 39.0% | | | | | 1.8% | | 87.5% | 10.7% | 30.5% | | | | 42.4% | 27.1% | | | | 10.2% | | 67.8% | 22.0% | | 30.5% | | 33.9% | 35.6% | | 35.6% | 37.3% | 27.1% |
| 3  N = 48 | | 0 | | 52.1% | 47.9% | 14.6% | | | | 45.8% | 39.6% | | | | | 2.3% | | 93.2% | 4.5% | 23.4% | | | | 48.9% | 27.7% | | | | 6.5% | | 71.7% | 21.7% | | 37.5% | | 29.2% | 33.3% | | 33.3% | 29.2% | 37.5% |
| 4  N = 62 | | 6.5% | | 46.8% | 46.8% | 19.4% | | | | 38.7% | 41.9% | | | | | 3.6% | | 87.5% | 8.9% | 29.0% | | | | 45.2% | 25.8% | | | | 1.7% | | 71.7% | 26.7% | | 33.9% | | 35.5% | 30.6% | | 45.2% | 24.2% | 30.6% |
| 5 Least  N = 83 | | 4.8% | | 54.2% | 41.0% | 19.3% | | | | 37.3% | 43.4% | | | | | 1.4% | | 91.9% | 6.8% | 22.9% | | | | 49.4% | 27.7% | | | | 6.1% | | 69.5% | 24.4% | | 36.1% | | 31.3% | 32.5% | | 36.1% | 33.7% | 30.1% |
| p value | | *.70* | | | | *.93* | | | | | | | | | | *.69* | | | | *.51* | | | | | | | | | *.69* | | | | | *.59* | | | | | *.65* | | |
| *Living arrangem.* | | *N = 296 Missing = 3* | | | | *N = 296 Missing = 3* | | | | | | | | | | | *N = 270 Missing = 29* | | | *N = 294 Missing = 5* | | | | | | | | | | *N = 290 Missing = 9* | | | | *N = 296 Missing = 3* | | | | *N = 296 Missing = 3* | | | |
| Live alone  N = 49 | | 4.1% | | 36.7% | 59.2% | 16.3% | | | | 38.8% | 44.9% | | | | | 4.4% | | 86.7% | 8.9% | 20.4% | | | | 42.9% | 36.7% | | | | 8.2% | | 67.3% | 24.5% | | 34.7% | | 30.6% | 34.7% | | 28.6% | 30.6% | 40.8% |
| Live with others  N = 247 | | 4.5% | | 53.8% | 41.7% | 17.0% | | | | 42.5% | 40.5% | | | | | 2.7% | | 90.2% | 7.1% | 26.9% | | | | 44.5% | 28.6% | | | | 5.0% | | 71.8% | 23.2% | | 32.4% | | 32.0% | 35.6% | | 40.5% | 29.6% | 30.0% |
| p value | | *.05* | | | | *.84* | | | | | | | | | | *.58* | | | | *.45* | | | | | | | | | *.64* | | | | | *.95* | | | | | *.22* | | |
| *Children in  household* | *N = 298 Missing = 1* | | | | | *N = 298 Missing = 1* | | | | | | *N = 272 Missing = 27* | | | | | | | | *N = 296 Missing = 3* | | | | | | *N = 292 Missing = 7* | | | | | | | | *N = 298 Missing = 1* | | | | *N = 298 Missing = 1* | | | |
| Children N = 118 | | 2.5% | | 50.0% | 47.5% | 15.3% | | | | 39.8% | 44.9% | | | | | 0.9% | | 93.7% | 5.4% | 22.0% | | | | 41.5% | 36.4% | | | | 6.1% | | 68.7% | 25.2% | | 33.9% | | 28.8% | 37.3% | | 41.5% | 25.4% | 33.1% |
| No children  N = 180 | | 5.6% | | 51.7% | 42.8% | 17.8% | | | | 43.3% | 38.9% | | | | | 4.3% | | 87.0% | 8.7% | 28.1% | | | | 46.6% | 25.3% | | | | 5.1% | | 72.3% | 22.6% | | 31.7% | | 33.9% | 34.4% | | 36.1% | 32.8% | 31.1% |
| p value | | *.39* | | | | *.58* | | | | | | | | | | *.15* | | | | *.11* | | | | | | | | | *.80* | | | | | *.66* | | | | | *.38* | | |
| *Contract type* | | *N = 294 Missing = 5* | | | | *N = 294 Missing = 5* | | | | | | | | | | *N = 269 Missing = 30* | | | | *N = 292 Missing = 7* | | | | | | | | | *N = 288 Missing = 11* | | | | | *N = 295 Missing = 4* | | | | | *N = 294 Missing = 5* | | |
| Permanent N =246 | | 4.1% | | 47.6% | 48.4% | 14.6% | | | | 41.5% | 43.9% | | | | | 2.7% | | 89.7% | 7.6% | 24.5% | | | | 43.3% | 32.2% | | | | 5.0% | | 71.8% | 23.2% | | 30.9% | | 32.5% | 36.6% | | 40.2% | 27.6% | 32.1% |
| Fixed term N = 20 | | 10.0% | | 60.0% | 30.0% | 45.0% | | | | 25.0% | 30.0% | | | | | 5.3% | | 89.5% | 5.3% | 45.0% | | | | 30.0% | 25.0% | | | | 10.5% | | 68.4% | 21.1% | | 55.0% | | 25.0% | 20.0% | | 45.0% | 35.0% | 20.0% |
| Atypical | | NA | | NA | NA | NA | | | | NA | NA | | | | | NA | | NA | NA | NA | | | | NA | NA | | | | NA | | NA | NA | | NA | | NA | NA | | NA | NA | NA |
| Self-employed/ Freelance  N = 28 | | 3.6% | | 71.4% | 25.0% | 17.9% | | | | 57.1% | 25.0% | | | | | 4.0% | | 88.0% | 8.0% | 22.2% | | | | 63.0% | 14.8% | | | | 7.1% | | 64.3% | 28.6% | | 35.7% | | 28.6% | 35.7% | | 21.4% | 39.3% | 39.3% |
| p value | | *.16* | | | | ***.01*** | | | | | | | | | | *.99* | | | | *.12* | | | | | | | | | *.82* | | | | | *.29* | | | | | *.05* | | |
| *Wage*  *precarity* | | *N = 243 Missing = 56* | | | | | *N = 243 Missing = 56* | | | | | | *N = 222 Missing = 77* | | | | | | | | *N = 241 Missing = 58* | | | | | | *N = 237 Missing = 62* | | | | | | | | *N = 243 Missing = 56* | | | *N = 243 Missing = 56* | | | |
| Low N = 93 | | 3.2% | | 52.7% | 44.1% | 11.8% | | | | 39.8% | 48.4% | | | | | 0 | | 92.1% | 7.9% | 22.6% | | | | 46.2% | 31.2% | | | | 5.4% | | 75.0% | 19.6% | | 29.0% | | 28.0% | 43.0% | | 31.2% | 29.0% | 39.8% |
| Moderate N = 106 | | 2.8% | | 50.0% | 47.2% | 17.9% | | | | 42.5% | 39.6% | | | | | 3.1% | | 91.7% | 5.2% | 26.9% | | | | 39.4% | 33.7% | | | | 3.9% | | 68.9% | 27.2% | | 34.9% | | 32.1% | 33.0% | | 41.5% | 32.1% | 26.4% |
| High N = 44 | | 6.8% | | 50.0% | 43.2% | 22.7% | | | | 45.5% | 31.8% | | | | | 2.7% | | 83.8% | 13.5% | 20.5% | | | | 47.7% | 31.8% | | | | 4.8% | | 76.2% | 19.0% | | 25.0% | | 40.9% | 34.1% | | 31.8% | 34.1% | 34.1% |
| p value | | *.80* | | | | *.31* | | | | | | | | | | *.25* | | | | *.83* | | | | | | | | | *.71* | | | | | *.38* | | | | | *.32* | | |
| *General  health* | | *N = 296 Missing = 3* | | | | | *N = 296 Missing = 3* | | | | | | *N = 270 Missing = 29* | | | | | | | | *N = 294 Missing = 5* | | | | | | *N = 290 Missing = 9* | | | | | | *N = 296 Missing = 3* | | | | | *N = 296 Missing = 3* | | | |
| Good N = 229 | | 3.9% | | 52.0% | 44.1% | 17.0% | | | | 45.4% | 37.6% | | | | | 2.4% | | 91.8% | 5.8% | 26.0% | | | | 50.2% | 23.8% | | | | 4.9% | | 73.2% | 21.9% | | 36.7% | | 34.5% | 28.8% | | 40.2% | 31.4% | 28.4% |
| Not good  N = 67 | | 4.5% | | 47.8% | 47.8% | 14.9% | | | | 31.3% | 53.7% | | | | | 4.8% | | 82.3% | 12.9% | 23.9% | | | | 25.4% | 50.7% | | | | 7.6% | | 63.6% | 28.8% | | 19.4% | | 23.9% | 56.7% | | 31.3% | 25.4% | 43.3% |
| p value | | *.75* | | | | *.05* | | | | | | | | | | *.07* | | | | ***<..0001*** | | | | | | | | | *.27* | | | | | ***<..001*** | | | | | *.07* | | |
| *Mental  well-being* | | *N = 295 Missing = 4* | | | | | | *N = 295 Missing = 4* | | | | | | *N = 270 Missing = 29* | | | | | | | | *N = 293 Missing = 6* | | | | | *N = 290 Missing = 9* | | | | | | | | *N = 295 Missing = 4* | | | *N = 295 Missing = 4* | | | |
| Low N = 34 | | 14.7% | | 14.7% | 70.6% | 17.6% | | | | 20.6% | 61.8% | | | | | 3.0% | | 84.8% | 12.1% | 17.6% | | | | 29.4% | 52.9% | | | | 0 | | 64.7% | 35.3% | | 26.5% | | 29.4% | 44.1% | | 32.4% | 20.6% | 47.1% |
| Average N = 261 | | 3.1% | | 55.2% | 41.8% | 16.9% | | | | 44.4% | 38.7% | | | | | 3.0% | | 90.7% | 6.3% | 26.6% | | | | 46.3% | 27.0% | | | | 6.3% | | 71.9% | 21.9% | | 33.3% | | 31.8% | 34.9% | | 39.5% | 30.7% | 29.9% |
| p value | | ***<..0001*** | | | | ***.02*** | | | | | | | | | | *.35* | | | | ***.01*** | | | | | | | | | *.10* | | | | | *.55* | | | | | *.12* | | |
| *Limiting  pre-existing  conditions* | | | *N = 287 Missing = 12* | | | | | | *N = 287 Missing = 12* | | | | | | *N = 261 Missing = 38* | | | | | | | | *N = 285 Missing = 14* | | | | | *N = 281 Missing = 18* | | | | | | | *N = 287 Missing = 12* | | | *N = 287 Missing = 12* | | | |
| Yes N = 54 | | 9.3% | | 42.6% | 48.1% | 22.2% | | | | 24.1% | 53.7% | | | | | 4.0% | | 90.0% | 6.0% | 30.2% | | | | 34.0% | 35.8% | | | | 1.9% | | 79.2% | 18.9% | | 20.4% | | 38.9% | 40.7% | | 33.3% | 24.1% | 42.6% |
| No N = 233 | | 3.0% | | 53.2% | 43.8% | 15.0% | | | | 47.2% | 37.8% | | | | | 2.8% | | 89.1% | 8.1% | 25.0% | | | | 47.0% | 28.0% | | | | 6.1% | | 69.3% | 24.6% | | 35.2% | | 30.5% | 34.3% | | 39.1% | 31.3% | 29.6% |
| p value | | *.07* | | | | ***.01*** | | | | | | | | | | *.78* | | | | *.23* | | | | | | | | | *.32* | | | | | *.11* | | | | | *.18* | | |

*Note. p values calculated with* Chi-square (χ^2^) *or Fisher’s exact tests. Modal factor level Ns included due to variation across health impacts.*

Table 2. Multivariate logistic regression model identifying independent predictors of positive and negative health impacts of home-working.

| *Factors* | *Health impacts of home-working* | | | | | | | | | | | | | |
| --- | --- | --- | --- | --- | --- | --- | --- | --- | --- | --- | --- | --- | --- | --- |
|  | *Feeling of loneliness (Valid N = 210)* | | *Mental well-being*  *(Valid N = 210)* | | *Smoking*  *(Valid N = 191)* | | *Eating well*  *(Valid N = 208)* | | *Drinking alcohol*  *(Valid N = 205)* | | *Exercise*  *(Valid N = 210)* | | *Work-life balance*  *(Valid N = 210)* | |
|  | Better | Worse | Better | Worse | Better | Worse | Better | Worse | Better | Worse | Better | Worse | Better | Worse |
| *Gender* |  |  |  |  |  |  |  |  |  |  |  |  |  |  |
| Men | *Reference* | | | | | | | | | | | | | |
| Women | *NA* | 0.97 [0.45-2.11]  p = .95 | 2.22 [0.68-7.19]  p = .19 | 1.32 [0.59-2.98]  p = .50 | *NA* | 2.16 [0.36-12.93]  p = .40 | 1.90 [0.71-5.10]  p = .20 | 1.23 [0.48-3.14]  p = .67 | 0.50 [0.06-4.24]  p = .53 | 0.73 [0.30-1.77]  p = .48 | 0.86 [0.34-2.18]  p = .74 | 0.76 [0.30-1.91]  p = .56 | 0.69 [0.28-1.69]  p = .41 | 0.94 [0.36-2.43]  p = .90 |
| *Age Group* |  |  |  |  |  |  |  |  |  |  |  |  |  |  |
| 18-29 Years | *NA* | 3.42 [0.86-13.62]  p = .08 | 0.54 [0.06-4.68]  p = .57 | 0.91 [0.22-3.85]  p = .90 | *NA* | 0.27 [0.01-10.45]  p = .49 | 2.28 [0.37-13.89]  p = .37 | 4.65 [0.80-27.18]  p = .09 | 82.69 [0.80-8520.16]  p = .06 | 3.04 [0.67-13.79]  p = .15 | 0.77 [0.14-4.22]  p = .78 | 2.51 [0.45-14.04]  p = .29 | 0.87 [0.17-4.49]  p = .87 | 1.18 [0.23-6.16]  p = .84 |
| 30-39 Years | *NA* | **3.32 [1.24-8.88]**  **p = .02** | 1.57 [0.39-6.29]  p = .52 | 1.83 [0.66-5.09]  p = .25 | *NA* | 4.38 [0.51-37.98]  p = .18 | 1.81 [0.53-6.20]  p = .34 | **4.56 [1.44-14.44]**  **p = .01** | 17.76 [0.60-528.57]  p = .10 | 1.65 [0.56-4.84]  p = .37 | 1.33 [0.41-4.35]  p = .64 | **6.10 [1.76-21.15]**  **p = .004** | 1.49 [0.46-4.84]  p = .51 | 2.75 [0.82-9.27]  p = .10 |
| 40-49 Years | *Reference* | | | | | | | | | | | | | |
| 50-59 Years | *NA* | 0.50 [0.19-1.31]  p = .16 | 0.61 [0.16-2.31] p = .46 | **0.30 [0.11-0.82]**  **p = .02** | *NA* | 1.19 [0.15-9.80]  p = .87 | 0.78 [0.26-2.36]  p = .66 | 0.45 [0.15-1.38]  p = .16 | 15.63 [0.42-581.83]  p = .14 | 1.27 [0.43-3.78]  p = .67 | 1.07 [0.37-3.08]  p = .90 | 2.05 [0.65-6.44]  p = .22 | 1.15 [0.39-3.34]  p = .80 | 0.77 [0.25-2.42]  p = .66 |
| 60-64 Years | *NA* | 1.47 [0.45 – 4.78]  p = .53 | 1.28 [0.26 – 6.39]  p = .76 | 0.62 [0.21-1.81]  p = .38 | *NA* | *NA* | 0.45 [0.10-1.93]  p = .28 | 0.36 [0.08-1.60]  p = .18 | 7.57 [0.13-446.45]  p = .33 | 0.92 [0.21-4.09]  p = .91 | 0.64 [0.17-2.40]  p = .51 | 1.43 [0.35-5.84]  p = .62 | 0.69 [0.18-2.65]  p = .59 | 0.31 [0.08-1.27]  p = .10 |
| *Deprivation Quintile* | | | | | | | | | | | | | |  |
| 1 Most deprived | *NA* | 1.16  [0.39-3.44]  p = .79 | 0.48 [0.09 – 2.46] p = .38 | 0.36 [0.12– 1.09] p = .07 | *NA* | *NA* | 2.14 [0.57-8.12]  p = .26 | 2.23 [0.64-7.82]  p = .21 | 0.75 [0.02-29.47]  p = .88 | 1.00 [0.31-3.24]  p = .99 | 1.80 [0.47-6.95]  p = .39 | 2.74 [0.78-9.64]  p = .12 | 3.12 [0.86-11.25]  p = .08 | 2.21 [0.59-8.33]  p = .24 |
| 2 | *NA* | 0.69  [0.28-1.73]  p = .43 | 0.72 [0.21 – 2.47] p = .60 | **0.32 [0.12– 0.86] p = .02** | *NA* | 1.00 [0.18-5.68]  p = 1.00 | 1.97 [0.70-5.52]  p = .20 | 0.58 [0.19-1.82]  p = .36 | 4.54 [0.29-70.87]  p = .28 | 0.85 [0.31-2.36]  p = .75 | 1.20 [0.42-3.45]  p = .73 | 1.19 [0.41-3.46]  p = .75 | 1.24 [0.46-3.34]  p = .67 | 0.53 [0.18-1.60]  p = .26 |
| 3 | *NA* | 0.94  [0.34-2.59]  p = .91 | 0.53  [0.13-2.20]  p = .38 | 0.57 [0.20– 1.65] p = .30 | *NA* | 0.04 [0.001-1.05]  p = .05 | 0.36 [0.08-1.53]  p = .17 | 0.71 [0.22-2.33]  p = .57 | *NA* | 0.37 [0.11-1.30]  p = .12 | 1.76 [0.56-5.52]  p = .34 | 1.73 [0.52-5.80]  p = .38 | 1.21 [0.38-3.91]  p = .75 | 1.72 [0.54-5.40]  p = .36 |
| 4 | *NA* | 1.38  [0.50-3.84]  p = .54 | 0.49  [0.12-2.05]  p = .33 | 0.62 [0.21–1.81] p = .38 | *NA* | 1.74 [0.27-11.37]  p = .57 | 1.45 [0.45-4.64]  p = .53 | 1.67 [0.52-5.38]  p = .39 | 0.38 [0.003-55.09]  p = .70 | 0.98 [0.32-2.99]  p = 0.97 | 1.51 [0.49-4.62]  p = .47 | 1.42 [0.43-4.69]  p = .56 | 3.15 [0.97-10.27]  p = .06 | 2.70 [0.76-9.58]  p = .12 |
| 5 Least deprived | *Reference* | | | | | | | | | | | | | |
| *Living arrangements* | | |  |  |  |  |  |  |  |  |  |  |  |  |
| Live alone | *NA* | 1.95  [0.75-5.06]  p = .17 | 2.11 [0.59-7.57]  p = .25 | 1.00 [0.36-2.72]  p = .99 | *NA* | 0.98 [0.12-8.12]  p = .99 | 0.74 [0.24-2.30]  p = .61 | 1.63 [0.53-4.98]  p = .40 | **40.73 [1.30-1271.66]**  **p = .04** | 1.61 [0.51-5.06]  p = .42 | 1.10 [0.39-3.16]  p = .85 | 1.05 [0.36-3.11]  p = .93 | 0.93 [0.32-2.74]  p = .90 | 1.44 [0.48-4.34]  p = .52 |
| Live with  others | *Reference* | | | | | | | | | | | | | |
| *Children in household* | | | | | | | | | | | | | | |
| Children | *NA* | 1.03  [0.43-2.45]  p = .95 | 0.95  [0.29-3.11]  p = .93 | 0.57 [0.23-1.39]  p = .21 | *NA* | 0.42 [0.06-3.12]  p = .39 | 0.78 [0.28-2.13]  p = .62 | 1.18 [0.43-3.26]  p = .75 | 7.70 [0.28-211.75]  p = .23 | 1.82 [0.69-4.79]  p = .23 | 1.37 [0.50-3.71]  p = .54 | 1.32 [0.47-3.71]  p = .59 | 1.65 [0.62-4.35]  p = .31 | 0.78 [0.28-2.18]  p = .64 |
| No children | *Reference* | | | | | | | | | | | | | |
| *Contract* |  |  |  |  |  |  |  |  |  |  |  |  |  |  |
| Permanent | *Reference* | | | | | | | | | | | | | |
| Fixed term | *NA* | **0.10**  **[0.02-0.61]**  **p = .01** | 5.30  [0.97-29.02]  p = .05 | 0.42  [0.07-2.58]  p = .35 | *NA* | 0.40 [0.03-6.32]  p = .52 | 1.59 [0.33-7.71]  p = .57 | 0.22 [0.03-1.43]  p = .11 | 0.51 [0.01-39.66]  p = .76 | 0.22 [0.04-1.33]  p = .10 | 3.49 [0.67-18.12]  p = .14 | 0.27 [0.03-2.61]  p = .26 | 0.84 [0.19-3.74]  p = .82 | 0.22 [0.03-1.59]  p = .13 |
| Atypical | *NA* | *NA* | *NA* | *NA* | *NA* | *NA* | *NA* | *NA* | *NA* | *NA* | *NA* | *NA* | *NA* | *NA* |
| Self-employed/ Freelance | *NA* | 0.38  [0.09-1.55]  p = .18 | 0.92  [0.19-4.39]  p = .92 | 0.26  [0.05-1.39]  p = .12 | *NA* | *NA* | 1.56 [0.41-6.01]  p = .52 | 0.21 [0.02-2.03]  p = .18 | 5.72 [0.15-218.04]  p = .35 | 0.81 [0.18-3.75]  p = .79 | 1.18 [0.31-4.56]  p = .81 | 1.07 [0.23-4.94]  p = .93 | 0.48 [0.13-1.82]  p = .28 | 0.61 [0.13-2.79]  p = .52 |
| *Wage precarity* |  |  |  |  |  |  |  |  |  |  |  |  |  |  |
| Low | *Reference* | | | | | | | | | | | | | |
| Moderate | *NA* | 0.92  [0.42-2.03]  p = .84 | 0.61  [0.19-1.97]  p = .41 | 0.57  [0.25-1.29]  p = .18 | *NA* | 0.65 [0.10-4.20]  p = .65 | 1.13 [0.44-2.88]  p = .80 | 1.06 [0.41-2.74]  p = .91 | 0.60 [0.03-11.51]  p = .74 | 2.04 [0.80-5.20]  p = .14 | 1.35 [0.53-3.43]  p = .53 | 0.58 [0.22-1.51]  p = .27 | 1.25 [0.50-3.10]  p = .64 | 0.68 [0.26-1.81]  p = .44 |
| High | *NA* | 0.92  [0.30-2.83]  p = .89 | 0.80  [0.18-3.59]  p = .77 | 0.45  [0.14-1.42]  p = .17 | *NA* | 4.12 [0.38-44.80]  p = .24 | 0.64 [0.17-2.42]  p = .51 | 1.01 [0.28-3.68]  p = .99 | 4.30 [0.05-352.92]  p = .52 | 1.35 [0.39-4.73]  p = .64 | 0.69 [0.20-2.42]  p = .56 | 0.37 [0.11-1.28]  p = .12 | 1.19 [0.35-4.04]  p = .78 | 1.08 [0.30-3.87]  p = .91 |
| *General health* |  |  |  |  |  |  |  |  |  |  |  |  |  |  |
| Good | *Reference* | | | | | | | | | | | | | |
| Not good | *NA* | 0.87  [0.35-2.16]  p = .77 | 0.87  [0.22-3.38]  p = .84 | 1.52  [0.60-3.84]  p = .38 | *NA* | **7.94 [1.03-61.43]**  **p = .047** | 2.49 [0.74-8.36]  p = .14 | **7.24 [2.33-22.49]**  **p = .001** | 4.54 [0.09-228.05]  p = .45 | **2.73 [1.05-7.10]**  **p = .04** | 0.56 [0.15-2.03]  p = .37 | **5.26 [1.72-16.12]**  **p = .004** | 0.74 [0.25-2.14]  p = .58 | 1.65 [0.56-4.85]  p = .36 |
| *Mental wellbeing* | | | | | | | | | | | | | | |
| Low | *NA* | **18.98**  **[3.53-102.07]**  **p = .001** | 1.03  [0.15-7.19]  p = .98 | **4.44**  **[1.25-15.79]**  **p = .02** | *NA* | 4.09 [0.48-35.00]  p = .20 | 1.43 [0.31-6.54]  p = .65 | 2.17 [0.57-8.23]  p = .26 | *NA* | 1.63 [0.51-5.23]  p = .42 | 0.71 [0.17-2.99]  p = .64 | 1.66 [0.47-5.86]  p = .43 | 0.82 [0.17-3.83]  p = .80 | 2.85 [0.72-11.25]  p = .13 |
| Average | *Reference* | | | | | | | | | | | | | |
| *Limiting pre-existing conditions* | | | | | | | | | | | | | | |
| Yes | *NA* | 1.96  [0.78-4.97]  p = .15 | 2.60  [0.65-10.40]  p = .18 | 2.60 [0.99-6.83]  p = .05 | *NA* | **0.08 [0.01-0.90]**  **p = .04** | 0.90 [0.30-2.68]  p = .85 | 0.61 [0.20-1.84]  p = .38 | *NA* | 0.50 [0.18-1.42]  p = .19 | 0.56 [0.19-1.68]  p = .30 | 0.35 [0.12-1.04]  p = .06 | 0.97 [0.34-2.77]  p = .95 | 1.28 [0.47-3.55]  p = .63 |
| No | *Reference* | | | | | | | | | | | | | |

*Note: Health outcome reference category is ‘no change’. Odds ratios adjusted for: gender, age, deprivation quintile, living arrangements, children in household, highest qualification level, contract type, wage precarity, job skill level, general health, mental well-being and limiting pre-existing conditions.*
