## Supplementary Material 5 for "Exploring the health impacts and inequalities of the new way of working: findings from a cross-sectional study"

### Supplementary Material 5: Preferences for future home-working

Table 1. Percentage of respondents indicating each preference for future home-working.

| *Factor* | *Preferences for future home-working* | | | | | *Significance^(1)^* |
| --- | --- | --- | --- | --- | --- | --- |
|  | *All working days* | *At least half* | *Less than half* | *No home working* | *Not sure* |  |
| *Gender* Missing N = 57 |  |  |  |  |  |  |
| Men  N = 198 | 11.1% | 24.2% | 18.7% | 19.2% | 26.8% | *p* = .12 |
| Women  N = 360 | 15.8% | 30.8% | 13.6% | 16.9% | 22.8% |  |
| *Age Group*  Missing N = 63 |  |  |  |  |  |  |
| 18-29 Years  N = 41 | 17.1% | 24.4% | 26.8% | 14.6% | 17.1% | *p* = .40 |
| 30-39 Years  N = 107 | 8.4% | 33.6% | 18.7% | 16.8% | 22.4% |  |
| 40-49 Years  N = 147 | 16.3% | 27.9% | 15.6% | 12.9% | 27.2% |  |
| 50-59 Years  N = 178 | 15.2% | 27.0% | 12.9% | 21.9% | 23.0% |  |
| 60-64 Years  N = 79 | 17.7% | 26.6% | 11.4% | 19.0% | 25.3% |  |
| *Deprivation Quintile* Missing N = 54 | | | | | | |
| 1 Most deprived  N = 106 | 10.4% | 24.5% | 12.3% | 23.6% | 29.2% | ***p* = .03** |
| 2  N = 133 | 13.5% | 24.8% | 12.0% | 16.5% | 33.1% |  |
| 3  N = 88 | 15.9% | 29.5% | 11.4% | 21.6% | 21.6% |  |
| 4  N = 102 | 16.7% | 36.3% | 20.6% | 9.8% | 16.7% |  |
| 5 Least deprived  N = 132 | 15.9% | 28.0% | 20.5% | 17.4% | 18.2% |  |
| *Living arrangements* Missing N = 56 | |  |  |  |  |  |
| Live alone  N = 110 | 10.9% | 22.7% | 14.5% | 24.5% | 27.3% | *p* = .13 |
| Live with others  N = 449 | 15.4% | 29.6% | 15.8% | 15.8% | 23.4% |  |
| *Children in Household*  Missing N = 54 | | | | | | |
| Children N = 203 | 15.3% | 31.5% | 15.8% | 14.8% | 22.7% | *p* = .55 |
| No children  N = 358 | 14.0% | 26.5% | 15.4% | 19.3% | 24.9% |  |
| *Contract type*  Missing N = 55 |  |  |  |  |  |  |
| Permanent N = 447 | 13.2% | 30.4% | 16.6% | 17.9% | 21.9% | ***p* = .004** |
| Fixed term  N = 33 | 15.2% | 33.3% | 18.2% | 15.2% | 18.2% |  |
| Atypical N = 24 | 8.3% | 20.8% | 0.0% | 20.8% | 50.0% |  |
| Self-employed/ Freelance  N = 56 | 26.8% | 12.5% | 12.5% | 14.3% | 33.9% |  |
| *Furlough* Missing N = 56 |  |  |  |  |  |  |
| Yes N = 105 | 11.4% | 22.9% | 12.4% | 16.2% | 37.1% | ***p* = .01** |
| No N = 454 | 15.2% | 29.7% | 16.1% | 18.1% | 20.9% |  |
| *Wage precariousness* Missing N = 136 | | | | | | |
| Low N = 145 | 13.8% | 28.3% | 17.2% | 24.1% | 16.6% | ***p* = .0001** |
| Moderate N = 194 | 11.3% | 36.1% | 15.5% | 14.9% | 22.2% |  |
| High N = 140 | 17.1% | 17.1% | 9.3% | 17.1% | 39.3% |  |
| *General health*  Missing N = 56 |  |  |  |  |  |  |
| Good N = 418 | 12.7% | 29.7% | 17.2% | 17.2% | 23.2% | *p* = .07 |
| Not good  N = 141 | 19.9% | 24.1% | 10.6% | 18.4% | 27.0% |  |
| *Mental wellbeing*  Missing N = 60 | | | | | | |
| Low N = 73 | 17.8% | 20.5% | 20.5% | 21.9% | 19.2% | *p* = .26 |
| Average N = 482 | 14.1% | 29.7% | 14.7% | 17.2% | 24.3% |  |
| *Limiting pre-existing conditions*  Missing N = 80 | | | | | | |
| Yes N = 119 | 19.3% | 23.5% | 13.4% | 18.5% | 25.2% | *p* = .31 |
| No N = 416 | 13.0% | 30.5% | 15.9% | 16.6% | 24.0% |  |

*Note: (1) p values calculated with* Chi-square (χ^2^) *or Fisher’s exact tests.*

Table 2. Multivariate multinomial regression model comparing preferences for future home-working across groups.

| *Factor* | *Preferences for future home-working* (Valid N = 435) | | | |
| --- | --- | --- | --- | --- |
|  | *All working days* | *Less than half* | *No home working* | *Not sure* |
| *Gender* |  |  |  |  |
| Men | *Reference* | | | |
| Women | **2.63 [1.06-6.50]**  **p = .04** | 0.57 [0.27-1.23]  p = .15 | 0.76 [0.38-1.51]  p = .43 | 0.63 [0.31-1.27]  p = .20 |
| *Age Group* |  |  |  |  |
| 18-29 Years | 0.94 [0.22-4.09]  p = .94 | 2.64 [0.72-9.67]  p = .14 | 0.99 [0.24-4.11]  p = .99 | 0.69 [0.17-2.84]  p = .61 |
| 30-39 Years | **0.29 [0.09-0.96]**  **p = .04** | 0.96 [0.35-2.61]  p = .94 | 0.99 [0.40-2.47]  p = .98 | 1.06 [0.44-2.57]  p = .89 |
| 40-49 Years | *Reference* | | | |
| 50-59 Years | 1.01 [0.38-2.66]  p = .99 | 1.04 [0.37-2.91]  p = .94 | 1.77 [0.71-4.39]  p = .22 | 1.00 [0.39-2.54]  p = 0.99 |
| 60-64 Years | 1.54  [0.47-5.10]  p = .48 | 0.54 [0.13-2.32]  p = .41 | 1.29 [0.42-4.02]  p = .66 | 0.62 [0.19-2.02]  p = .43 |
| *Deprivation Quintile* | | | | |
| 1 Most deprived | 1.03  [0.34-3.16]  p = .96 | 0.35 [0.10-1.16]  p = .09 | 1.09 [0.42-2.82]  p = .86 | 1.34 [0.48-3.69]  p = .58 |
| 2 | 1.26  [0.47-3.36]  p = .65 | 0.49 [0.18-1.33]  p = .16 | 0.76 [0.31-1.88]  p = .56 | 1.26 [0.50-3.21]  p = .63 |
| 3 | 0.88  [0.29-2.65]  p = .82 | 0.35 [0.11-1.17] p = .09 | 1.13 [0.44-2.87]  p = .80 | 0.57 [0.19-1.73]  p = .32 |
| 4 | 0.72  [0.24-2.15]  p = .55 | 0.88 [0.33-2.32]  p = .79 | **0.27 [0.08-0.89]**  **p = .03** | 0.98 [0.35-2.76]  p = .97 |
| 5 Least deprived | *Reference* | | | |
| *Living arrangements* | |  |  |  |
| Live alone | 1.38  [0.53-3.57]  p = .51 | 1.20  [0.45-3.20]  p = .72 | **2.40 [1.05-5.50]**  **p = .04** | 1.99 [0.85-4.66]  p = .12 |
| Live with others | *Reference* | | | |
| *Children in household* | | | | |
| Children | 1.10 [0.44-2.72]  p = .84 | 1.05  [0.44-2.54]  p = .91 | 1.40 [0.63-3.15]  p = .41 | 1.03 [0.46-2.33]  p = .94 |
| No children | *Reference* | | | |
| *Contract type* |  |  |  |  |
| Permanent | *Reference* | | | |
| Fixed term | 1.98  [0.42-9.35]  p = .39 | 0.41  [0.07-2.35]  p = .32 | 1.18 [0.30-4.64]  p = .82 | 1.13 [0.30-4.32]  p = .86 |
| Atypical | 0.76  [0.11-5.47]  p = .79 | NA | 0.43 [0.07-2.81]  p = .38 | 1.19 [0.28-5.09]  p = .81 |
| Self-employed/ Freelance | **6.98**  **[1.98-24.59]**  **p = .002** | 1.09  [0.22-5.39] p = .92 | 0.50 [0.09-2.91]  p = .44 | 2.80 [0.81-9.64]  p = .10 |
| *Furlough* |  |  |  |  |
| Yes | 0.82  [0.28-2.39]  p = .72 | 1.41  [0.53-3.74]  p = .49 | 1.80 [0.76-4.26]  p = .18 | **3.09 [1.40-6.82]**  **p = .01** |
| No | *Reference* | | | |
| *Wage precarity* | | | | |
| Low | *Reference* | | | |
| Moderate | 0.54  [0.23-1.28]  p = .16 | 1.10  [0.49-2.49]  p = .82 | 0.55 [0.26-1.15]  p = .11 | 0.98 [0.43-2.24]  p = .97 |
| High | 1.27  [0.48-3.36]  p = .63 | 1.52  [0.51-4.49]  p = .45 | 1.23 [0.48-3.13]  p = .67 | **4.32 [1.69-11.05]**  **p = .002** |
| *General health* |  |  |  |  |
| Good | *Reference* | | | |
| Not good | 1.64 [0.66-4.09]  p = .29 | 0.51  [0.18-1.43]  p = .20 | 1.30 [0.57-2.98]  p = .53 | 0.95 [0.39-2.30]  p = .91 |
| *Mental wellbeing* |  |  |  |  |
| Low | 0.90  [0.30-2.67]  p = .85 | 1.44 [0.45-4.62]  p = .54 | 0.67 [0.24-1.89]  p = .45 | 0.42 [0.14-1.24]  p = .12 |
| Average | *Reference* | | | |
| *Limiting pre-existing conditions* | | | | |
| Yes | 1.41  [0.57-3.50]  p = .45 | 1.61  [0.61-4.23]  p = .34 | 1.18 [0.51-2.77]  p = .70 | 1.11 [0.47-2.61]  p = .82 |
| No | *Reference* | | | |

*Note: Reference category for preferences was the most populated selection – working from home on half or more of all working days.* *Odds ratios adjusted for: gender, age, deprivation quintile, living arrangements, children in household, highest qualification level, contract type, furlough, wage precarity, job skill level, general health, mental well-being and limiting pre-existing conditions.*
